# Race, class, and place modify mortality rates for the top 12 causes of death, 1999-2021

**DOI:** 10.1101/2022.06.14.22276404

**Authors:** Allison Formanack, Ayush Doshi, Rupa Valdez, Ishan Williams, J Randall Moorman, Pavel Chernyavskiy

## Abstract

**Objectives:** To disarticulate the associations of race (whiteness), class (socioeconomic status), and place (county) with risk of cause-specific death in the US.

**Methods:** We studied mortality in US counties for 11 causes of death (1999-2019) and COVID-19 (2020-2021). We adjusted for race and age using the American Community Survey and socioeconomic status using the Area Deprivation Index. Bayesian regressions with spatial county effects were estimated for inference.

**Results:** County whiteness and socioeconomic status modified death rates; geospatial effects differed by cause of death. Other factors equal, a 20% increase in county whiteness was associated with 5-8% increase in death from three causes and 4-15% reduction in death from others, including COVID-19. Other factors equal, advantaged counties had significantly lower death rates, even when juxtaposed with disadvantaged ones. Geospatial patterns of residual risk varied by cause of death. For example, cancer and heart disease death rates were better explained by age, socioeconomic status, and county whiteness than were COVID-19 and suicide deaths.

**Conclusions:** There are important independent contributions from race, class, and geography to risk of death in the US.

## INTRODUCTION

### Motivation

Disparity and inequity in healthcare are legion in the United States (US). The National Institutes of Health defines levels of analysis in health disparities to be environmental, sociocultural, behavioral, and/or biological. Among these categories, public health literature is typically focused on geographical and socio-economic factors, racial and ethnic factors, health behaviors, as well as physiological factors, such as the prevalence and type of disabilities. Further, formally-recognized priority populations for disparities research include, but are not limited to: racial, ethnic, sexual orientation and gender minorities, socio-economically disadvantaged populations, rural populations, and disabled populations^1^. Unfortunately, validated public data with an appropriate level of geographic detail that contain measurement on these factors and sub-populations are limited. However, racial and ethnic composition (*race and ethnicity*), socio-economic status (*class*), and geographic location (*place*) have better established measures and validated data sources, and thus best lend themselves to further study and will be the focus of this article.

These three variables - self-reported race and ethnicity, class, and place - are well-known risk factors for death, alongside, of course, age. Even high-quality recent efforts to investigate US health disparities frequently consider one or two of these at a time, rather than adjusting simultaneously for all three^2–5^. In cases where all three are considered, researchers tend to focus on one or only on a few causes of death^6,7^. Thus, less is known about how these three fundamental drivers of health disparities will contribute jointly to mortality, nor is it known how these drivers differ by cause of death.

Not only do we expect all three of race, class, and place to matter, but we also expect interaction among them. For example, due to systemic racism and structural housing discrimination, racial minorities tend to live in more disadvantaged communities across the US^8^. Intuitively, both joint and independent contributions of the three factors are not expected to be identical for all causes of death. For example, it is not clear that the variables associated with health disparity and geospatial landscape are the same for suicide as they are for influenza. Here, we use a county-level regression framework to investigate how socioeconomic status, county racial make-up, and geography contribute to risk of age-adjusted mortality for the top-12 causes of death in 2020. This approach allows us to analyze the contribution of each factor when adjusted for the others.

### Race and ethnicity, class, and place

Race and ethnicity are social and cultural constructs that are not biologically based, but nevertheless impact death rates. Our focus is on the racial make-up of US counties, and our measure is the proportion of residents that identify as non-Hispanic white in the 5-year American Community Survey. We call this metric the “whiteness” of the county, following critical race theorists who distinguish between “white people” as a descriptor and “whiteness” as a sociocultural construct and lived experience of systemic racial privilege and advantage^9–11^. In the US, white raciality provides structural and systemic advantages that affect health outcomes, even as socio-cultural definitions of whiteness have shifted over time^12^. As a consequence of racial privilege and structural racism, non-Hispanic whiteness is often regarded as normative in relation to all other racial identities, in particular Black or African American populations^13^. Our research objective is to determine the association of non-Hispanic whiteness, with mortality rates of the top-12 causes of death, a list that prominently includes COVID-19. By focusing on whiteness defined in this way, our analysis directly measures the effects of privilege associated with whiteness on health outcomes.

Class, by which we here mean socioeconomic status (SES), also affects health outcomes but is harder to measure. We used the 2018 Area Deprivation Index (ADI) percentiles for US block-groups that were aggregated to the level of county^14^. The ADI is a validated factor-based index composed of 17 census variables that measure areal levels of education, employment, income, poverty, and housing^15^. By spatializing these factors, the ADI is an effective predictor for mortality^15,16^ and life expectancy^16,17^. Worse ADI^18^ and rising food insecurity^19^ are associated with cardiovascular mortality, and that high ADI is a risk factor for hospital readmission of the same magnitude as Chronic Obstructive Pulmonary Disease (COPD) and more than diabetes^20^. This is especially true for sepsis^21^ and after surgery^22–25^. Importantly, we note that the formulation of the ADI omits racial and ethnic makeup, but may well encode its effects^9–11^.

Place, like race/ethnicity and SES, has an impact on health outcomes. The integral role of place (i.e., spatial context) in health disparities is formally recognized by the US Department of Health and Human Services^26,27^ via its Social Determinants of Health (SDH). As such, counties reflect the essential variability in socio-demographics and the built environment, and thereby serve as effective proxies for the SDH domains of Health Care Access and Quality, Neighborhood and Built Environment, and Social and Community Context. In addition, risk factors for common diseases cluster: for example, hypertension is more common in the southern US, and thus we can expect higher rates of death from heart and kidney disease. Most recently, the pandemic has thrown into especially sharp relief the differences in death rates across the country. Likewise, COVID vaccination uptake varies regionally^28^, as do local pandemic-related community prevention efforts, leading to strong geospatial patterns of pandemic mortality^29^.

Our analysis of place-based risk bears elaboration. Here, we consider the patterns of ADI formed by adjacent counties in addition to the risk associated with residence in each county. To do so, we partition US counties into one of several geographic profiles: clusters of advantage and disadvantage, outliers of advantage and disadvantage, and neither clustered nor outlying counties. As such, our analysis formally reflects the (dis)advantage of each county relative to its neighbors, which could reflect factors otherwise not accounted for by analyzing each county individually. For example, we might expect geographical differences in access to healthcare, socio-cultural resources, and so on.

Here, we bring race, class, and place into the same analytic frame to measure their contributions and interactions^30^ in influencing population health outcomes. Our statistical models also reveal the impact of latent variables - we call them *geographic risk modifiers* - that capture the joint impact of all factors not explicitly given by our measures of race, class, and place. We expect these factors to be complex and nuanced, and plausible mechanisms must draw on concepts of syndemics and intersectionality.

## METHODS

We obtained death certificate-based county level counts and person-years at risk for the top 11 causes of death 1999-2019 from the CDC Wonder database^31^, and for COVID-19 over 2020-2021 from the Johns Hopkins Center for Systems Science and Engineering^32^. COVID-19 mortality spanned January 1, 2020 - October 17, 2021. Due to missing ADI, we analyzed data in all but 7 counties, 4 of which were in Hawaii. We show the unadjusted and unsmoothed mortality rates, depicted as cause-specific Z-scores in Supplementary Figure 1. Mapped in this manner, a county with a value of 2 for heart disease, for example, has an unadjusted rate two standard deviations above the US average for heart disease.

### ADI Clustering using Local Moran’s I

To concentrate the effects of areal deprivation and reduce confounding with the smooth spatial county effects, we employed Local Moran’s I^33^ computed using GeoDa open-source software^34^. This analysis effectively assigns each county to either a deprivation cluster or outlier status based on its Area Deprivation Index (ADI) and the ADI of its immediate neighbors. The resultant disadvantaged clusters were disadvantaged counties surrounded by disadvantage, whereas advantaged clusters were advantaged counties surrounded by advantage. Disadvantaged outliers and advantaged outliers were spatial outliers that are statistically significantly different from their neighbors. We used alpha=0.05 to detect clusters and outliers. Together with the non-cluster-non-outlier grouping - where Local Moran’s *I* yielded p-values > 0.05 - we created a 5-level categorical variable that jointly reflects the levels of deprivation of each county, as well as that of its first-order neighbors.

### Statistical Analysis

We estimated 12 Bayesian regression models - one per cause of death - each with a person-years-at-risk “offset term” (*E*_*i*_) and explanatory variables of: county age demographics (% aged <18, % aged >65) and % non-Hispanic white residents from the 5-year American Community Survey^35^, as well as the 5-level ADI cluster type (reference = non-cluster-non-outliers). Each model also included a cause-specific spatial county effect (*u*_*i*_) to account for spatial correlation, which was specified using the recent BYM2 formulation^36^. We used the negative-binomial likelihood that accounts for overdispersion in the data. The analysis was implemented using standard code within the brms package version 2.15 in R^37^.

The expected count of cause-specific deaths for the *i*th county was specified as:

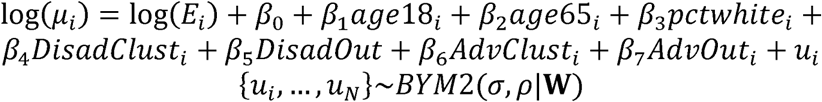

where **W** is the county adjacency matrix. Counties were deemed adjacent using the “queen” rule: if their borders shared at least one vertex. For causes of death where exact mortality counts were censored by the CDC due to being too small, we applied an identically-specified left-censored negative binomial regression^38^.

Spatial effects {*u*_*i*_, …, *u*_*N*_ } reflect patterns of risk attributable to geographic location, after adjustments due to age, race, and class. In other words, they reflect the aggregate contribution of unmeasured or unmeasurable factors unique to each county. In the results, we present estimates on a common Relative Risk (RR) scale for all causes of death, motivating their interpretation as a *geographic risk modifier*, where RR = 1 reflects no modification beyond what is explained by the other explanatory variables.

All quantitative explanatory variables were centered and scaled prior to analysis. We adjusted the prior distributions and MCMC sampling parameters in each model to balance execution times - typically long in spatial analysis - with convergence metrics. This sampling strategy was adjusted until convergence was deemed acceptable for each cause of death. The details of our sampling parameters, as well as the prior distributions used, are shown in Supplementary Table 2.

As a sensitivity analysis, we used standard 5-year categorical age groups instead of continuous age percentages, as well as the more parsimonious Intrinsic Conditional Autoregressive correlation structure. Using categorical ages resulted in lower uncertainty due to pseudo-replication of ADI cluster effects across age groups, but neither the direction nor the statistical significance of the coefficients were substantially altered. Similarly, using a more parsimonious correlation structure did not qualitatively alter our findings. Thus, we report the results from analyses with the simplified age structure and using the BYM2 spatial model, as shown above.

## RESULTS

### Race: County whiteness predicted mortality rates

Greater county whiteness was associated with lower risk of death for 9 of 12 leading causes, after adjusting for age, class, and place (Figure 1). Counties with a higher proportion of white residents had at least 4% lower rates of 8 of the top 12 causes of death. The greatest disadvantages occurred in kidney disease, septicemia, diabetes, and COVID-19, where death rates were 12 to 15% lower in counties with a higher proportion of white residents. Deaths due to suicide, chronic lower respiratory disease, and Alzheimer’s disease were higher in whiter counties. The adjusted risk of death due to cancer, on the other hand, was less affected by county whiteness with Relative Risk (RR)=0.99 (95% Credible Interval 0.98, 0.99) per 20% increase in county whiteness.

**Figure 1.**
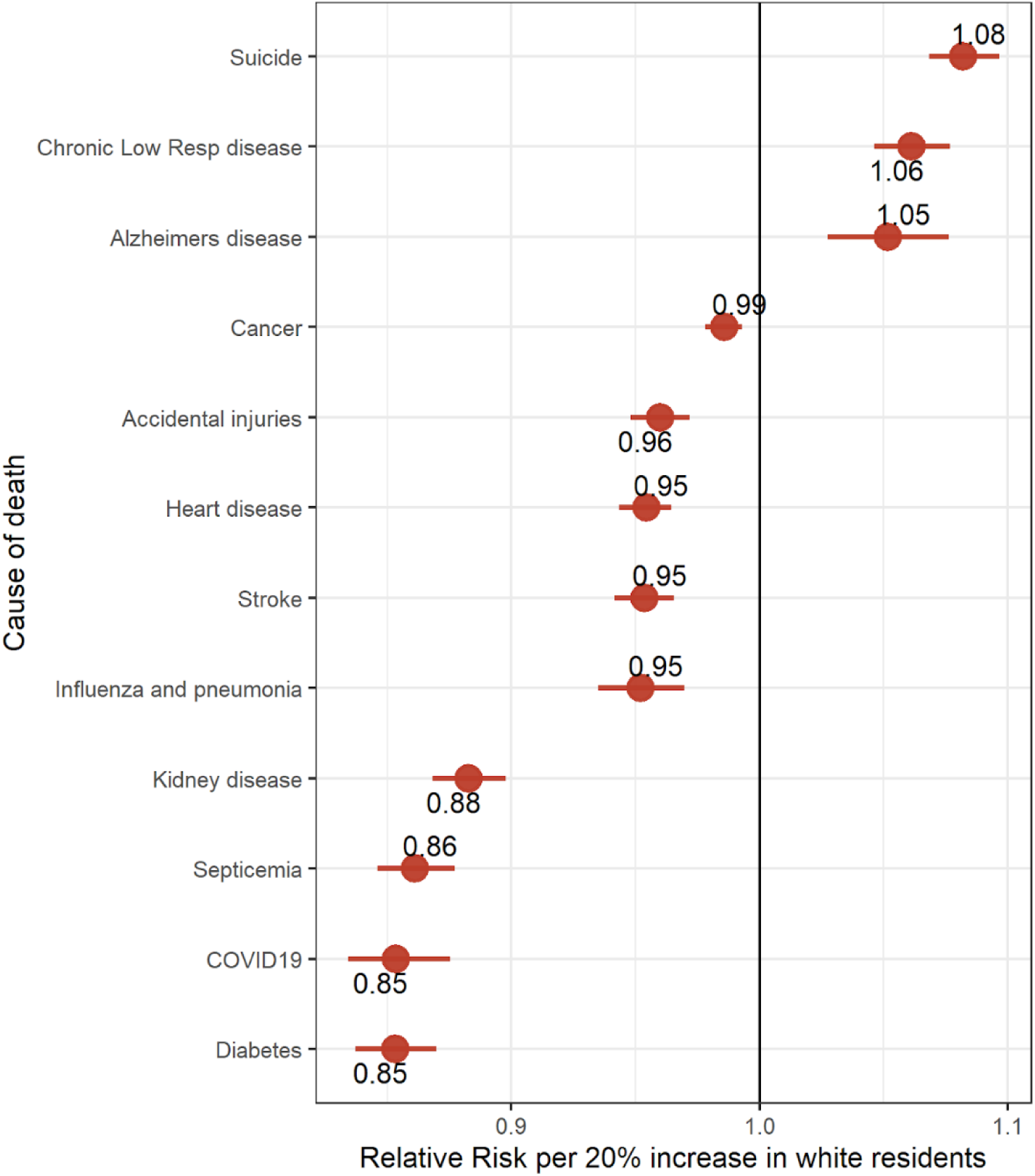
Race and cause of death. The data points are cause-specific adjusted relative risks (RR) per 20% (1 SD) increase in county whiteness (% of non-Hispanic white residents), sorted according to the RR estimate. Bars are 95% equal-tail Credible Intervals. All RR are adjusted for age, class (Area Deprivation Index clusters), and place (cause-specific geographic risk modifiers); RR = 1 indicates that the risk is independent of race (county whiteness).

### Class: Area Deprivation Index clusters predicted mortality rates

We found that 696 (22.2%) counties were in a disadvantaged cluster, 504 counties (16.1%) were in an advantaged cluster, 21 counties (0.7%) were disadvantaged outliers, 28 counties (0.9%) were advantaged outliers, and the remaining counties (60.1%) were neither outliers nor in a cluster. Advantaged-cluster counties were generally situated in states along the Pacific coast, in the Northeast metropolitan corridor and New England, and in portions of the Mountain West states (Colorado, Wyoming, Montana, and Utah). Disadvantaged-cluster counties were concentrated in non-coastal counties in the South, portions of the Midwest, and also in portions of Appalachia. An interactive online map of the computed ADI clusters and outliers can be accessed using this link.

Residence in an advantaged cluster was associated with significantly lower risk for 9 of 12 causes of death. Relative to non-cluster-non-outlier counties, these Relative Risks (RR) ranged from 0.89 (95% CrI: 0.80, 0.99) for COVID-19 to 0.98 (0.96, 1.00) for cancer (Figure 2; bottom panel). Conversely, residence in a disadvantaged cluster was associated with significantly higher risk for 8 of 12 causes of death, with RRs ranging from 1.02 (1.01, 1.04) for cancer to 1.11 (1.07, 1.15) for influenza and pneumonia (Figure 2; top panel).

**Figure 2.**
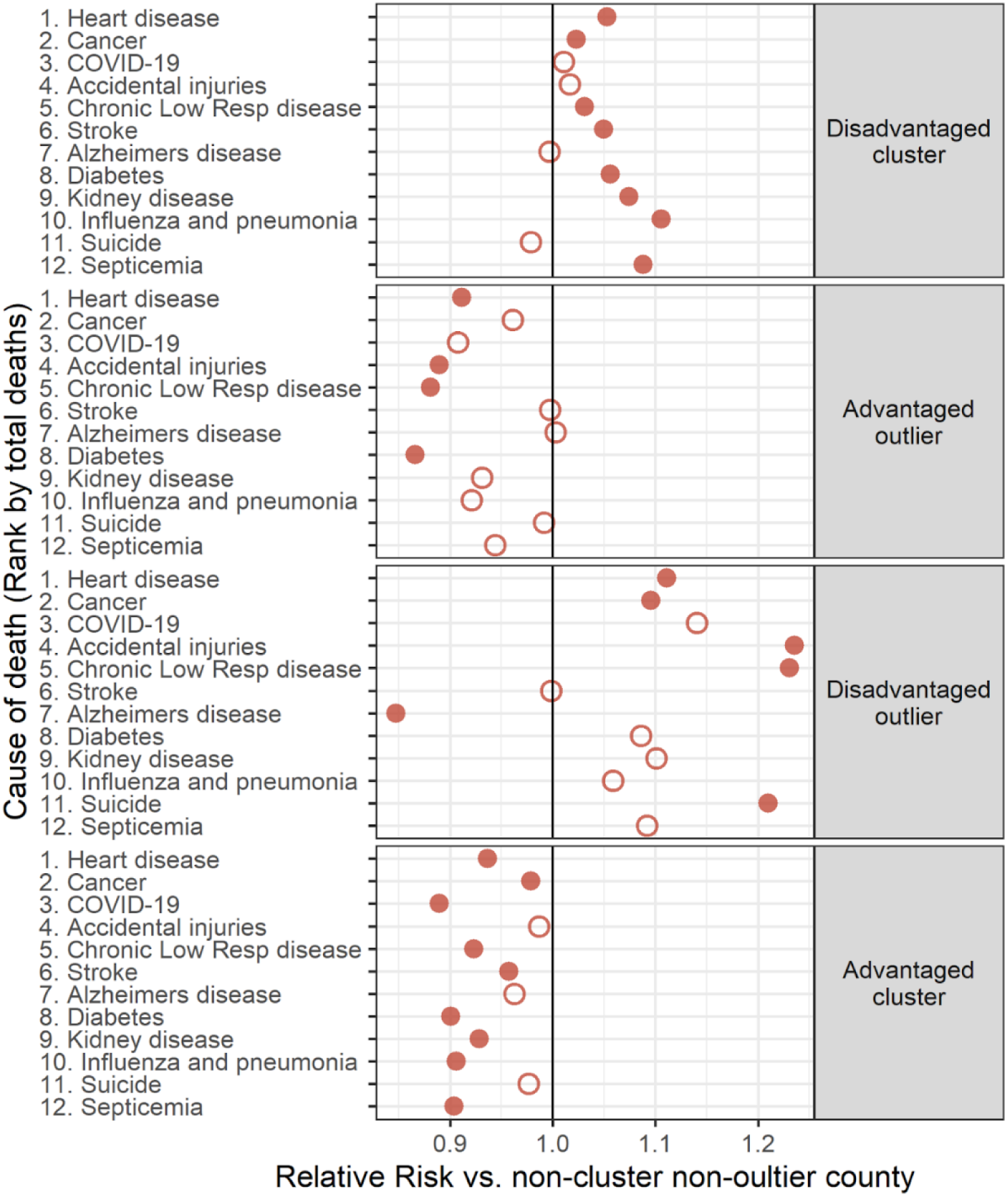
Class and cause of death. Points are cause-specific adjusted relative risks (RR) for counties belonging to an ADI-based disadvantaged cluster, advantaged outlier, disadvantaged outlier, or advantaged cluster *vs*. non-cluster-non-outlier. Open circles indicate that RR=1 is included in a 95% equal-tail Credible Interval; RR=1 indicates risk of death is independent of cluster type. All RR are adjusted for age, race and ethnicity (whiteness), and place (cause-specific geographic risk modifiers) and are ranked by the total number of deaths within each panel.

Residence in an outlier county was often associated with more extreme RR compared to residence in a cluster. For example, the RR for chronic lower respiratory disease in a disadvantaged outlier was 19.4% higher than a disadvantaged cluster. On the other hand, the RR for Alzheimer’s disease in an advantaged outlier was 4.7% lower than in an advantaged cluster.

Surprisingly, Alzheimer’s disease mortality was unlike all other causes of death with an RR substantially below 1 in disadvantaged outlier counties and not significantly different elsewhere.

### Place: geographic risk modifiers differed markedly by cause of death

Figure 3 shows that geographic risk modifiers for kidney disease, septicemia, and COVID-19 were elevated in the Southeast and Northeast, and for COVID-19 across parts of the South, Midwest, and the Mountain-West states. On the other hand, geographic risk modifiers for accidental deaths and suicides tended to be higher across the West, including Alaska. Modifiers for Alzheimer’s further differed and were localized in several counties within South Dakota, North Dakota, and Washington. Finally, modifiers for chronic lower respiratory disease were localized in Colorado and Kentucky. To our surprise, the top two causes of death - heart disease and cancer - had the least variable effects, with little geographic modification of risk due after accounting for other factors.

**Figure 3.**
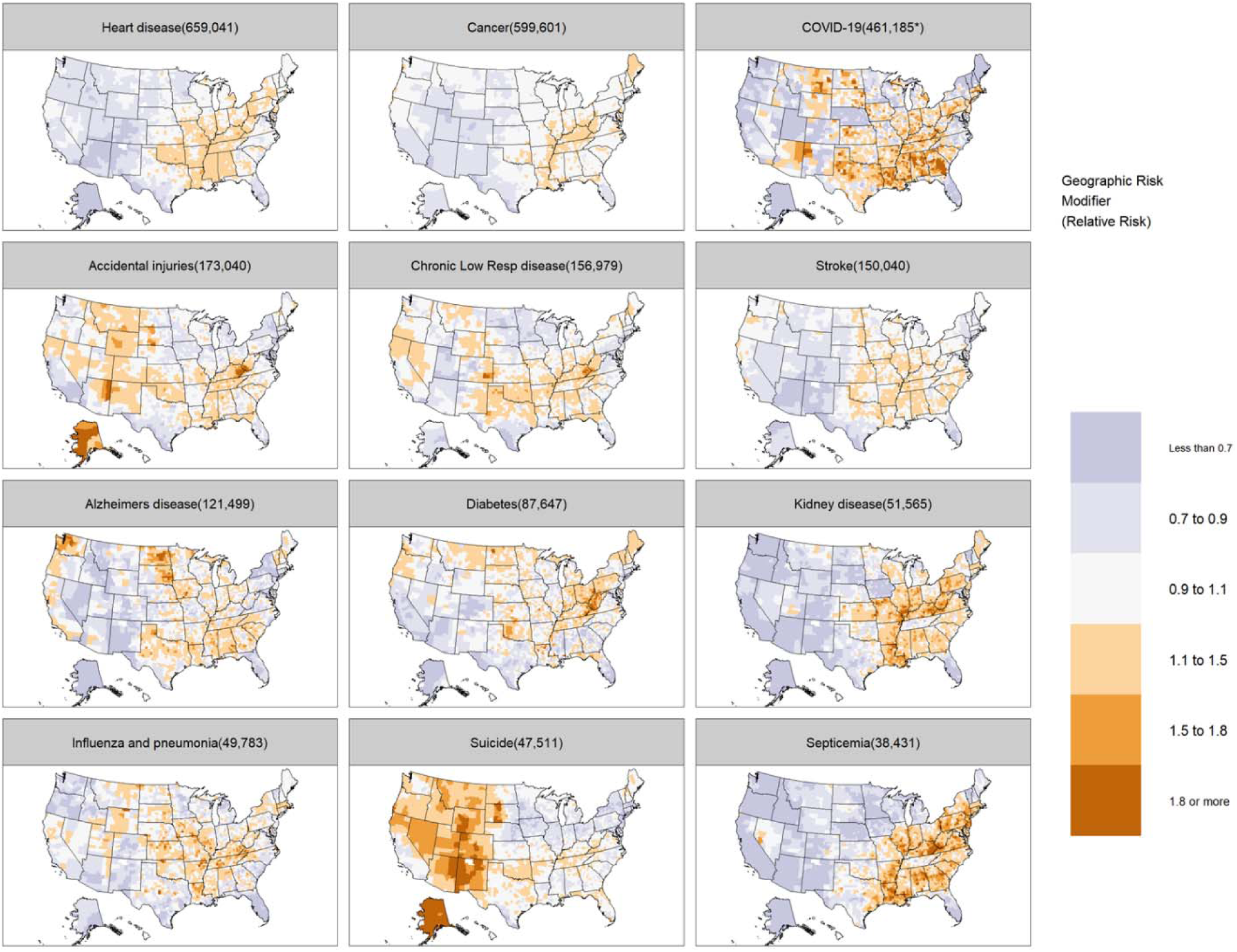
Place and cause of death. Maps reflect cause-specific geographic risk modifiers after adjustments for age, race and ethnicity (whiteness), and class (ADI-based clusters). The color scale reports the Relative Risk and can be interpreted as a place-based multiplier of cause-specific mortality.

Geographic risk modifiers for heart disease, cancer, kidney disease, and septicemia were inter-correlated (Spearman *ρ* > 0.65), and thus co-located in a similar set of counties (Supplementary Figure 2). Similarly, risk modifiers for stroke and heart disease (*ρ* = 0.61), and accidental injuries and suicide (*ρ* = 0.56) co-located. Conversely, risk modifiers for suicide were statistically significantly anti-correlated with heart disease (*ρ* = -0.15), stroke (*ρ* = -0.18), and COVID-19 (*ρ* = -0.37), indicating those occur in different sets of counties.

The high correlation between cause-specific geographic risk modifiers and observed mortality rates indicates important non-race, non-class risk factors were present. Spearman correlations ranged from 0.88 (COVID-19) to 0.39 (cancer) (Supplementary Table 1). At the extremes, these correlations indicate cancer mortality patterns were much better explained by age, race, and class than patterns of COVID-19 mortality.

## DISCUSSION

For the leading causes of death in 2020, we jointly evaluated the impacts of self-reported race and ethnicity, class, and place at the county level. The major strengths of our study include the use of sophisticated analytical tools, authoritative data sources, and the complete list of the 12 most common causes of death. Moreover, we treated these fundamental risk factors as statistically independent, measuring the influence of one adjusted for the others. Our measures of race/ethnicity, class, and place do not overlap; for example, our measure of class (the ADI) does not include race, ethnicity, or place. Our major finding is that race, class, and place each contribute to mortality risk and these contributions differ markedly by cause of death. Our findings suggest there are at least three mechanisms responsible for structural inequalities present in US healthcare today.

Race and ethnicity, measured here by county whiteness, was protective for 9 causes and not protective for 3 causes of death. Whiteness affords no physiological benefits, but it historically affords social advantages that allow access to healthcare and other mechanisms for improved outcomes. For example, although the protective whiteness effect for heart disease - the number one cause of death - was relatively small among the causes studied, treatments have long been administered to fewer Black patients than white^39^. The protective whiteness effect for COVID-19 - the third leading cause in 2020 - was particularly pronounced in our data, which mirrors previous findings of greater risk to ethnic and racial minority populations^6,7,40^. From a statistical perspective, working with county whiteness affords us sufficient numeric range to maximize statistical power to detect disparities when they are present. In fact, county-level data on every possible racial and ethnic subgroup are sparse and estimates may lead to spurious conclusions.

Class, measured by the ADI, was associated with risk of death from all of the top 12 causes of death. As expected, we found that disadvantaged counties had worse outcomes and advantaged counties had better outcomes. To our surprise, we found that juxtaposition of counties with different levels of advantage did not have intermediate effects. That is, disadvantaged outlier counties did not benefit from the proximity of advantaged neighbors, nor did advantaged outlier counties suffer from disadvantaged neighbors. It is plausible that counties are geographically too large for these kinds of intermediate effects to arise; our work should be reproduced on smaller geographic units (e.g., census tracts) to determine if the patterns we observed persist. Additionally, there were relatively few ADI outlier counties (49 across both types of outliers), and we found no consistent features of these counties. We note, however, that 5 of the 28 advantaged outlier counties housed large research universities in the southern US, 4 of them in Mississippi alone. Within these counties, outcomes were much better than for their immediate neighbors. The mechanisms are not known but might include the presence of a stable workforce with healthcare benefits, as well as superior health and community infrastructure, for example: access to specialist physicians, community investment in greenspaces, and access to healthy foods.

Place, after adjusting for race and class, had a strong effect on mortality through geographic risk modifiers that differed substantially by cause of death. We observed patterns consistent with known clusters, such as the “Heart Failure belt” and “Stroke belt” in the Southeast^41,42^, the “Suicide belt” in the West^43^, and to a lesser degree, “Cancer alley” in counties along the Mississippi River in the South^44,45^. Our analysis also indicates clusters of COPD in Colorado and Kentucky, perhaps explained by the history of mining, as evidenced by the presence of federal Black Lung clinics^46^. Additionally, our results highlight previously unreported regional patterns, such as the clustering of high-risk geographic modifiers for kidney disease and septicemia in the eastern US, notably excluding Tennessee and Florida.

We note especially the effects of race, class, and place on COVID-19 deaths. Race had a profound effect: county whiteness protected against COVID deaths in this analysis, consistent with other reports. The effect was one of the strongest that we found and was matched only by diabetes mortality. Likewise, class had a strong effect: residents of advantaged clusters were most protected from COVID deaths among the twelve causes under study. Additionally, with respect to place, we found complex nation-wide patterns of COVID-19 deaths. For example, several Native American nations in Arizona, New Mexico, Wyoming, and Montana have mortality risk in excess of what can be attributed to race and class, highlighting the local challenges of healthcare delivery in those locales^47^. In stark contrast, large portions of the US West, and especially California, Oregon, Washington, Utah, and Nevada, were relatively protected from COVID-19 mortality, after adjusting for race and class.

While our analysis statistically isolates each variable from the others, we note the potential intersections of race, class, and place for some of the causes of death. For example, in areas that benefit from sustained economic growth (e.g., “Silicon Valley”), which may form advantaged clusters and outliers, we find favorable mortality for diabetes and chronic lower respiratory disease. Likewise, we find that class and race may intersect for Alzheimer’s disease, which is not only of higher prevalence among African American/Black populations and underdiagnosed as well^48^. Finally, we find that place and race intersect in the western US, with higher county whiteness as well as rates of suicide^49^ and other “deaths of despair”^50^.

Taken together, we regard these intersections of race, class, and place as empirical evidence that supports syndemics theory. As a conceptual framework, *syndemics* treats health inequities as a confluence of biosocial factors, in particular the structural, historical, and cultural systems that inform individual decision-making^51^. Syndemics analyses have been done for each cause of death we identify in this paper, including COVID-19^52,53^; however, there is little evidence that supports syndemics theories in practice^54^. Here, we present an empirical basis for syndemics that shows where spatially-structured risk accentuates socially-structured risk. Specifically, we are able to demonstrate: (1) how whiteness affects mortality for the top 12 causes of death; (2) how socioeconomic advantage and disadvantage is mediated by racial and geographic proximity; and (3) that morbidities cluster in specific biosocial patterns independent of race and class. We argue that these conclusions cannot be reached without simultaneously attending to the intersection of race, class, and place as syndemic risk modifiers.

This work represents the first known evidence^55,56^ for syndemics theory that does not rely on behavioral data; however, we acknowledge several limitations. Using aggregated data carries an inherent risk of committing ecological bias. To this end, we emphasize that our smallest unit of analysis is the county and take care to limit our scope of inference no further. Counts of deaths due to COVID have been subject to underreporting^57^, especially for racial and ethnic minorities. Thus, we are limited by the quality of the data publicly available through the CDC. Because we focus our article on race, class, and place, several potential risk factors were not part of this analysis, perhaps most prominently, biological sex. Indeed, there are known sex differences for several leading causes of deaths, including heart disease^58^, cancer^59^, and suicide^60^. Likewise, we do not consider other factors that contribute to health disparities, such as the prevalence and type of disabilities^61,62^ in the population, or the prevalence of individuals who identify as something other than cisgender and heterosexual^63^. However, investigating the interaction of these factors with race, class, and place falls outside the scope of the current work, and presents a fecund direction for future research. Finally, we note that county whiteness is a function of self-reported race, ascertained at the county level through the American Community Survey. And, while we report non-Hispanic white as the reference standard, we note that the reality of racial and ethnic makeup in the US is far more complex than a simple “white/non-white” binary.

## PUBLIC HEALTH IMPLICATIONS

We offer two perspectives on public health implications. First, our article supports syndemics theory, in that we offer empirical evidence for race, class, and place as intersectional, rather than independent operators affecting differential outcomes in mortality for the top 12 causes. Second, existing measures of deprivation like the ADI are useful for identifying macro-level trends, but their usefulness is limited for local health departments, which tend to operate at the county level, with a decentralized structure meant to respond to local health conditions. In addition to other findings, our study illustrates how macro-level datasets can be analyzed in a way that is more suited to the needs of local health departments and their partners.

## Data Availability

Data available upon reasonable request to authors.

## ACKNOWLEDGEMENTS

We thank D.S. Wessel, MD, for help with literature review and K. Krahn for assistance with manuscript preparation.

## Notes

### Competing Interest Statement

The authors have declared no competing interest.

### Funding Statement

This study did not receive any funding.

### Summary of Updates

Introduction, Methods, Discussion text revised. Citations added throughout manuscript.

