## Supplementary for "Race, class, and place modify mortality rates for the top 12 causes of death, 1999-2021"

**SUPPLEMENTARY MATERIALS**

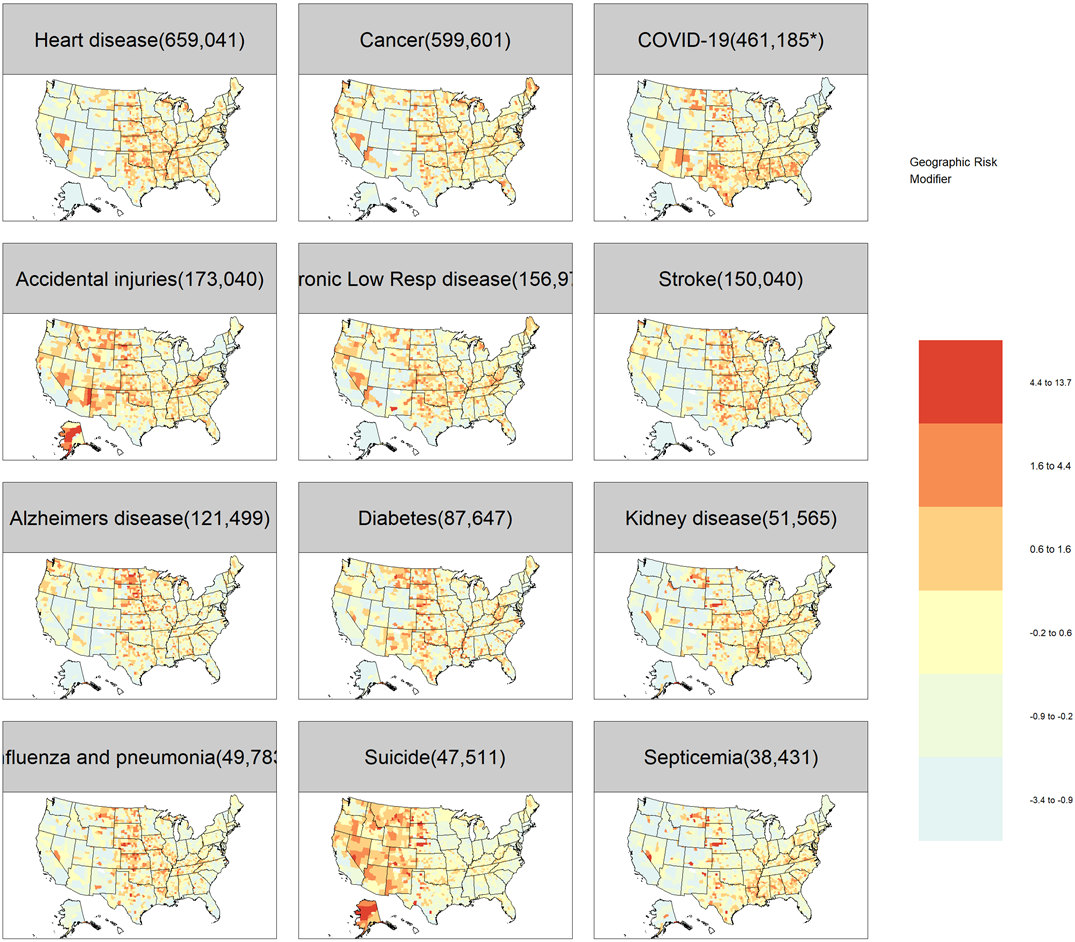

Supplementary Figure 1. Unadjusted and unsmoothed mortality rates shown as Z-scores, standardized to the cause-specific mean mortality rate. The color scale corresponds to the number of standard deviations each county lies above/below the cause-specific mean mortality. By design, these data are not geographically smoothed; thus, readers are urged to apply care in interpreting extreme Z-scores in sparsely populated counties.

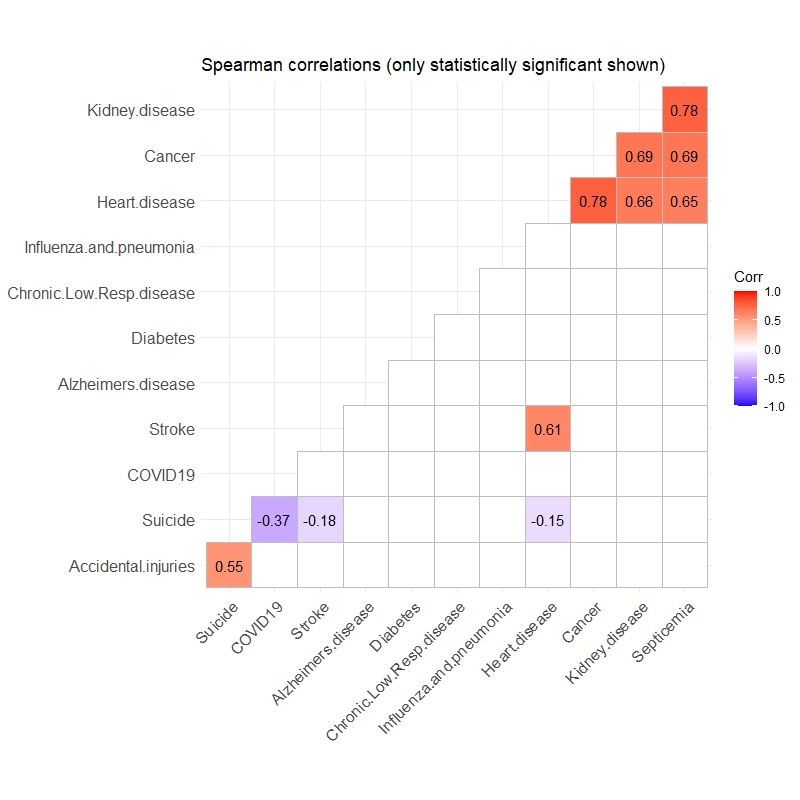

Supplementary Figure 2. Correlation among geographic risk modifiers (i.e., latent spatial effects) for each cause of death, adjusted for age, race, and class. Spearman correlations were used to capture possible non-linear relationships. Empty squares indicate correlation was not statistically significant at 95% confidence level. A clustering algorithm was applied to visually group like correlations together, as is apparent for kidney disease, cancer, heart disease, and septicemia, for example.

| Cause of death with ranking by total deaths | Spearman correlation of spatial latent effects with observed mortality | Standard Deviation of log-spatial latent effects (95% Credible Interval) |
| --- | --- | --- |
| 1. Heart disease | 0.592 | 0.75 (0.68, 0.82) |
| 2. Cancer | 0.399 | 0.47 (0.53, 0.51) |
| 3. COVID-19 | 0.884 | 2.17 (2.07, 2.26) |
| 4. Accidental injuries | 0.757 | 0.98 (0.93, 1.04) |
| 5. Chronic Lower Respiratory disease | 0.675 | 1.01 (0.93, 1.10) |
| 6. Stroke | 0.635 | 0.78 (0.70, 0.86) |
| 7. Alzheimer’s disease | 0.782 | 1.43 (1.28, 1.59) |
| 8. Diabetes | 0.709 | 1.23 (1.12, 1.36) |
| 9. Kidney disease | 0.709 | 1.33 (1.25, 1.41) |
| 10. Influenza and pneumonia | 0.756 | 1.23 (1.12, 1.35) |
| 11. Suicide | 0.798 | 0.94 (0.89, 0.99) |
| 12. Septicemia | 0.734 | 1.42 (1.34, 1.50) |

Supplementary Table 1. Summary metrics of geographic risk modifiers (i.e. spatial latent effects) for each cause of death, estimated via the BYM2 model after adjusting for race, class, and place. High correlation between risk modifiers and observed mortality indicates that the latent effects reflect an important spatial process and the explanatory variables in the model are relatively poor predictors of cause-specific mortality. Spatial smoothing parameters close to 1 indicate strong spatial correlation among the latent effects; spatial smoothing parameters close to 0 indicate latent effects are spatially independent.

| Cause of death with ranking by total deaths | Largest R-hat | Smallest Effective Sample Size | Divergent transitions (% of post-warmup) | Warm-up samples (total samples) per chain | Adapt delta parameter (Max treedepth) | Prior distributions used |
| --- | --- | --- | --- | --- | --- | --- |
| 1. Heart disease | 1.01 | 449 | 0 (0) | 500 (3500) | 0.99 (15) | 1; ~N+(0,2.5); ~B(1,1); ~N+(0,40) |
| 2. Cancer | 1.01 | 586 | 0 (0) | 500 (3500) | 0.99 (15) | 1; ~N+(0,2.5); ~B(1,1); ~N+(0,40) |
| 3. COVID-19 | 1.10 | 12 | 0 (0) | 300 (1500) | 0.90 (10) | ~N(0, 2.5); ~N+(0,1.5); ~B(0.5,0.5); ~G(0.01,0.01) |
| 4. Accidental injuries | 1.01 | 363 | 0 (0) | 500 (3500) | 0.99 (15) | 1; ~N+(0,2.5); ~B(1,1); ~N+(0,40) |
| 5. Chronic Lower Respiratory disease | 1.01 | 316 | 0 (0) | 500 (3500) | 0.99 (15) | 1; ~N+(0,2.5); ~B(1,1); ~N+(0,40) |
| 6. Stroke | 1.01 | 179 | 0 (0) | 500 (3500) | 0.99 (15) | 1; ~N+(0,2.5); ~B(1,1); ~N+(0,40) |
| 7. Alzheimer’s disease | 1.16 | 19 | 13 (0.11) | 500 (3500) | 0.99 (15) | 1; ~N+(0,2.5); ~B(1,1); ~N+(0,40) |
| 8. Diabetes | 1.06 | 67 | 0 (0) | 500 (3500) | 0.99 (15) | 1; ~N+(0,2.5); ~B(1,1); ~N+(0,40) |
| 9. Kidney disease | 1.02 | 293 | 0 (0) | 500 (3500) | 0.99 (15) | 1; ~N+(0,2.5); ~B(1,1); ~N+(0,40) |
| 10. Influenza and pneumonia | 1.05 | 97 | 0 (0) | 500 (3500) | 0.99 (15) | 1; ~N+(0,2.5); ~B(1,1); ~N+(0,40) |
| 11. Suicide | 1.00 | 680 | 0 (0) | 500 (3500) | 0.99 (15) | 1; ~N+(0,2.5); ~B(1,1); ~N+(0,40) |
| 12. Septicemia | 1.00 | 298 | 0 (0) | 500 (3500) | 0.99 (15) | 1; ~N+(0,2.5); ~B(1,1); ~N+(0,40) |

Supplementary Table 2. Bayesian Markov Chain Monte Carlo (MCMC) implementation details for the top 12 causes of deaths. No-U-Turn Hamiltonian Monte Carlo via the brms package with 4 parallel chains was used for all estimation. All models underwent visual inspection of MCMC chains and parameter space to ensure satisfactory convergence occurred.
